# Fecal pH as a marker of stunting among children hospitalized for diarrhea and other non-diarrheal pathologies

**DOI:** 10.1101/2025.08.26.25334519

**Authors:** Md. Shabab Hossain, Md Mahbubul Hoque, Mustafa Mahfuz, Tahmeed Ahmed

## Abstract

**Background:** Fecal pH is a simple, non-invasive diagnostic tool used for initial screening of certain gastrointestinal (GI) diseases. Increased fecal pH indicates reduction of beneficial microbiota in the gut, which has emerged as a key factor contributing to stunting. The purpose of this study was to investigate the association of fecal pH with stunting in hospitalized children.

**Methods:** This cross-sectional study was conducted on 200 children aged 06-24 months getting admitted in icddr,b Dhaka Hospital with diarrhea and Dhaka Shishu Hospital for other non-diarrheal pathologies. Length-for-age Z scores (LAZ) was measured and data on factors affecting linear growth was recorded. Fecal pH was measured on freshly collected stool samples following standard procedure. Multivariate linear regression was performed to explore relationship between fecal pH and LAZ scores.

**Results:** The mean fecal pH of diarrheal and non-diarrheal children was 5.54±0.98 and 5.95±0.76, respectively. After inclusion of factors affecting linear growth into the regression model, a statistically significant inverse association between fecal pH and LAZ scores was observed in non-diarrheal children (p<0.01). However, such association did not apply for diarrheal children.

**Conclusion:** Increased fecal pH in non-diarrheal children was found to have significant association with stunted growth, making fecal pH a possible indirect determinant of childhood stunting. However, no such associations were observed in case of diarrheal children.

## 1 Introduction

Fecal pH is a simple, non-invasive, as well as inexpensive diagnostic tool that is being used for the initial screening of certain gastrointestinal (GI) diseases for decades. For instance, while acidic stool indicates *E*.*coli* and rotaviral diarrhea, amoebic dysentery, and severe acute malnutrition (SAM) often accompanied by carbohydrate (CHO) intolerance (1, 2); alkaline stool is associated with colitis and colonic malignancy (3, 4). It is also applied as a tool for assessing prognosis of systemic pathologies in critically ill hospitalized patients where elevated fecal pH indicates bacteremia and increased mortality (5). Studies also found strong association of fecal pH with gut microbiota status where a marked change in the infant gut microbiome was reflected by an altered fecal pH (6, 7).

At present, one-third of children in developing countries are affected by chronic malnutrition or stunting (8), which ultimately results in a reduced neuro-developmental potential and poor cognitive function of the children (9). Along with several other factors that are responsible for childhood stunting, recent studies suggesting the role of altered gut microbiota status or dysbiosis as an important factor contributing to this process has generated the idea of exploring indirect, non-invasive methods to explore the gut microbiota status (10–14). A pilot trial on non-diarrhoeal stunted and at risk of stunting community children enrolled from the Bangladesh Environmental Enteric Dysfunction (BEED) study was recently conducted in a slum of Dhaka, Bangladesh to explore the relation between fecal pH and linear growth (15). The results showed fecal pH to have a significantly inverse association with LAZs of these children (15).

Elevated fecal pH indicates a profound reduction of *Bifidobacterium* (6), the most important gut symbiont in infants and the abundance of which is indicative of an optimum linear growth (6, 13, 14). This species is particularly important because of its unique ability to consume human milk oligosachharides (HMOs), which are prebiotic oligosaccharides only present in human milk. HMOs promote growth of nonpathogenic microflora and prevent pathogenic microorganisms from adhesion to the host epithelium, and thus provides protection against infections, diarrhea (16), and other morbidities which ultimately leads to under-nutrition and stunting. Short chain fatty acids (SCFAs) are produced as a byproduct of the fermentation process that results from the presence of *Bifidobacteria* as well as some other commensals like *Clostridium* clusters XIVa and IV in the gut. As evident from the name, SCFAs are acidic in nature and have a significant effect on fecal pH (6). *Clostridium* clusters XIVa and IV has also been found to be reduced in critically ill hospitalized patients, whose fecal pH were also found to be elevated (17).

Among these SCFAs acetate, propionate, and butyrate are the most abundant (18, 19). Studies comparing conventional and gnotobiotic animals showed that large intestinal SCFA concentrations were increased manifolds by the presence of microbiota (20), while all of these three primary SCFAs were virtually undetectable in gnotobiotic animals (21). Butyrate lowers colonic pH contributing to acidification of stool (7). It is calorie-rich and its chronic depletion contributes to under-nutrition and stunting (22). Butyrate also prevents the colonization of facultative anaerobic enteropathogens and its reduction promotes overrepresentation of aerobic pathogens which results in increased GI diseases, again leading to under-nutrition and stunting (22). The main butyrate-producing bacteria are *Clostridia*, especially clusters IV and XIVa (19), while the production of other SCFAs like acetate and proprionate is mediated by *Bifidobacteria* (23). Therefore, along with *Bifidobacteria*, elevated fecal pH is also associated with reduction in these butyrate producing commensal *Clostridium* clusters (24), the reduction of which are also considered as a signature of stunting (22).

Figure 1 shows a schematic diagram of the possible mechanism referred here.

**Figure 1.**
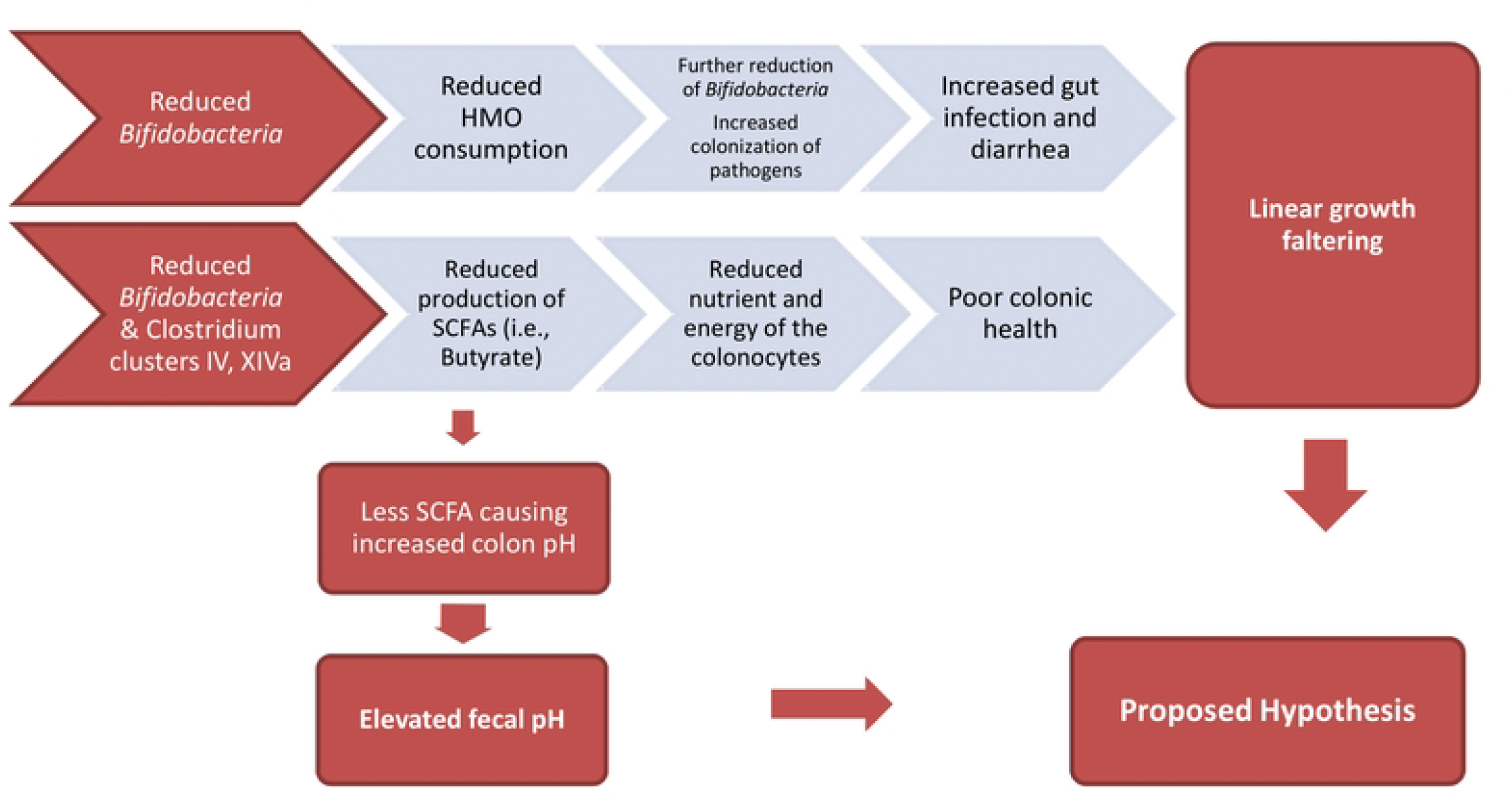
Schematic diagram of possible patho-physiology

Therefore, an indirect inverse relationship exists between fecal pH and linear growth, reflecting the gut microbiota status, the values of which however might differ in diarrheal and non-diarrheal states. We therefore hypothesized fecal pH to be a marker of child growth, and sought to evaluate this association both in diarrheal and non-diarrheal hospitalized children.

## Methods

### Study site and Data collection

This study, cross-sectional in design, was conducted on 200 children aged between 06-24 months getting admitted in icddr,b Dhaka Hospital with diarrhea and Dhaka Shishu Hospital for other non-diarrheal pathologies. The first participant was enrolled on 18 August, 2019 and the participant recruitment was completed on 23 February, 2020. Diarrhea was defined as the passage of three or more loose or liquid stools per day. The diarrheal children were further categorized into two groups: one with severe acute malnutrition (SAM) and the other without it. 100 children with diarrhea, of them 50 having a WLZ score below −3, with or without nutritional edema were enrolled from the icddr,b Dhaka Hospital. Another 100 non-SAM children admitted in Dhaka Shishu Hospital were enrolled as the non-diarrheal group. The diarrheal group consisted of children aged 6-24 months, with passage of three or more loose or liquid stools per day and a WLZ score below −3 and/or MUAC<11.5 cm, with or without nutritional edema for SAM cohort. While, non-diarrhoeal, non-SAM children referred to children aged 6-24 months, with WLZ > −1 (using WHO growth standards 2006) and no evidence of diarrhea.

A written informed consent was collected from a parent or guardian on behalf of the participants. Anthropometric, socio-demographic and food frequency questionnaire (FFQ) data were recorded using hard copy questionnaires and 20-25g of stool sample was collected for assessing the fecal pH. Children with any congenital disorder or deformity, severe sepsis or septic shock, and any known history of chronic illness, presence of severe anemia, tuberculosis, other chronic diseases or any congenital disorder or deformity were excluded from the study.

Weight and length of the participants was assessed using standard guidelines (25). Socio-economic, morbidity, dietary intake and breastfeeding data was collected using hard copy questionnaires on enrolment (26, 27). Qualitative food frequency questionnaires (FFQ) was used for the assessment of dietary intake (26, 27). Ethical approval was taken from Ethical Review Committee (ERC) of the Institutional Review Board (IRB) of icddr,b. Approved protocol number is PR-19074.

### Measuring stool pH

A portable pH meter, which yielded quantitative results, was used to measure the fecal pH. Following standard operating procedures (6, 28, 29), 20-25 grams of fresh fecal samples was collected immediately after defecation and 2-3 grams of fecal sample was kept in a sterile specimen container.10 ml de-ionized water was mixed and homogenized with this fecal specimen. The probes of the device were then submerged into this mixture and retained for one minute and the reading was recorded. The whole process was performed within first 120 minutes of sample collection. A portable stool pH meter from Hanna Instruments, USA, was used for the process.

### Data Analysis

Statistical analyses were performed using SPSS version 20.0 (IBM). Means and standard deviation (SD) of means or medians and inter-quartile ranges (IQR), wherever applicable, were used to describe the distribution. Correlation between fecal pH and LAZ scores was done using Pearson correlation and scatter plots for individual cohorts. Fecal pH is significantly associated with certain diarrheal conditions, ie. *E*.*coli* and rotaviral diarrhea, amoebic dysentery, and severe acute malnutrition (SAM) accompanied by carbohydrate (CHO) intolerance (2, 30). Therefore, data analyses were performed separately on diarrheal and non-diarrheal children. As linear growth measured in LAZ score is the outcome variable, at first bivariate linear regression was performed between LAZ scores with each individual factors followed by multivariate linear regression to assess the association between fecal pH and LAZ scores after adjusting for relevant factors that also included incorporation of several aspects of socio-economic status: access to improved **W**ater and sanitation, the selected approach to measuring household wealth (**A**ssets), **M**aternal education, and **I**ncome (i.e. the WAMI index) (31).

## Results

A total of 200 children (100 diarrheal, 100 non-diarrheal) with median age of 12±6 months underwent fecal pH examination. Among them, 137 (68%) were male and 63 (32%) were female. The mean fecal pH of diarrheal and non-diarrheal children was 5.54±0.98 and 5.95±0.76, respectively, while the mean LAZ of diarrheal and non-diarrheal children was −1.77±1.26 and − 0.99±1.07, respectively. Table 1 shows the socio-demographic characteristics of the study population.

**Table 1.** Demographic characteristics and results of the respondents.

| Characteristics | Diarrheal cohort<br>Mean ± SD*<br>(n=100) | Non-diarrheal cohort<br>Mean ± SD*<br>(n=100) | p-value |
| --- | --- | --- | --- |
| Mean age of children (Months) | 11.76 ± 4.16 | 13.32±4.12 | 0.53 |
| Male, % | 67% | 70% | 0.65 |
| Fecal pH | 5.54±0.98 | 5.95±0.76 | 0.13 |
| Length for age Z-score (LAZ) | -1.77±1.26 | -0.99±1.07 | 0.47 |
| Weight for age Z-score (WAZ) | -2.61±1.52 | -0.66±0.99 | 0.24 |
| Weight for height z-score (WHZ) | -2.32±1.48 | -0.28±0.92 | 0.19 |
| Mother's age (Years) | 34.9 ± 8.8 | 34.3±18.9 | 0.58 |
| Mother’s schooling years | 8.78 ± 3.6 | 6±3.6 | 0.17 |
| Mother’s currently working status |  |  |  |
| Yes | 14% | 9% | 0.44 |
| No | 86% | 91% |  |
| Primary cause of hospitalization, % |  |  |  |
| Diarrhea | 96% | 0% | 0.00 |
| ARI*/Pneumonia | 0% | 31% |  |
| Febrile illness | 2% | 43% |  |
| Others | 2% | 26% |  |
| Mother’s hand wash practice |  |  |  |
| Wash hand with soap after helping child defecate, % |  |  |  |
| Always | 73% | 78% | 0.13 |
| Sometimes | 23% | 18% |  |
| Rarely | 0% | 3% |  |
| Never | 4% | 1% |  |
| Monthly family income in USD*, median (IQR*) | 150 (100, 248) | 200 (150, 300) | 0.06 |

\* SD= Standard deviation; \*ARI= Acute Respiratory Tract Illness; USD= United States Dollars; IQR=Inter-quartile range

Pearson correlation between fecal pH and LAZ scores of the non-diarrheal children showed a statistically significant inverse correlation (−0.21, p<0.01). Scatter plot also showed a similar trend between stool pH and the LAZ scores (Figure-3).

**Figure 2.**
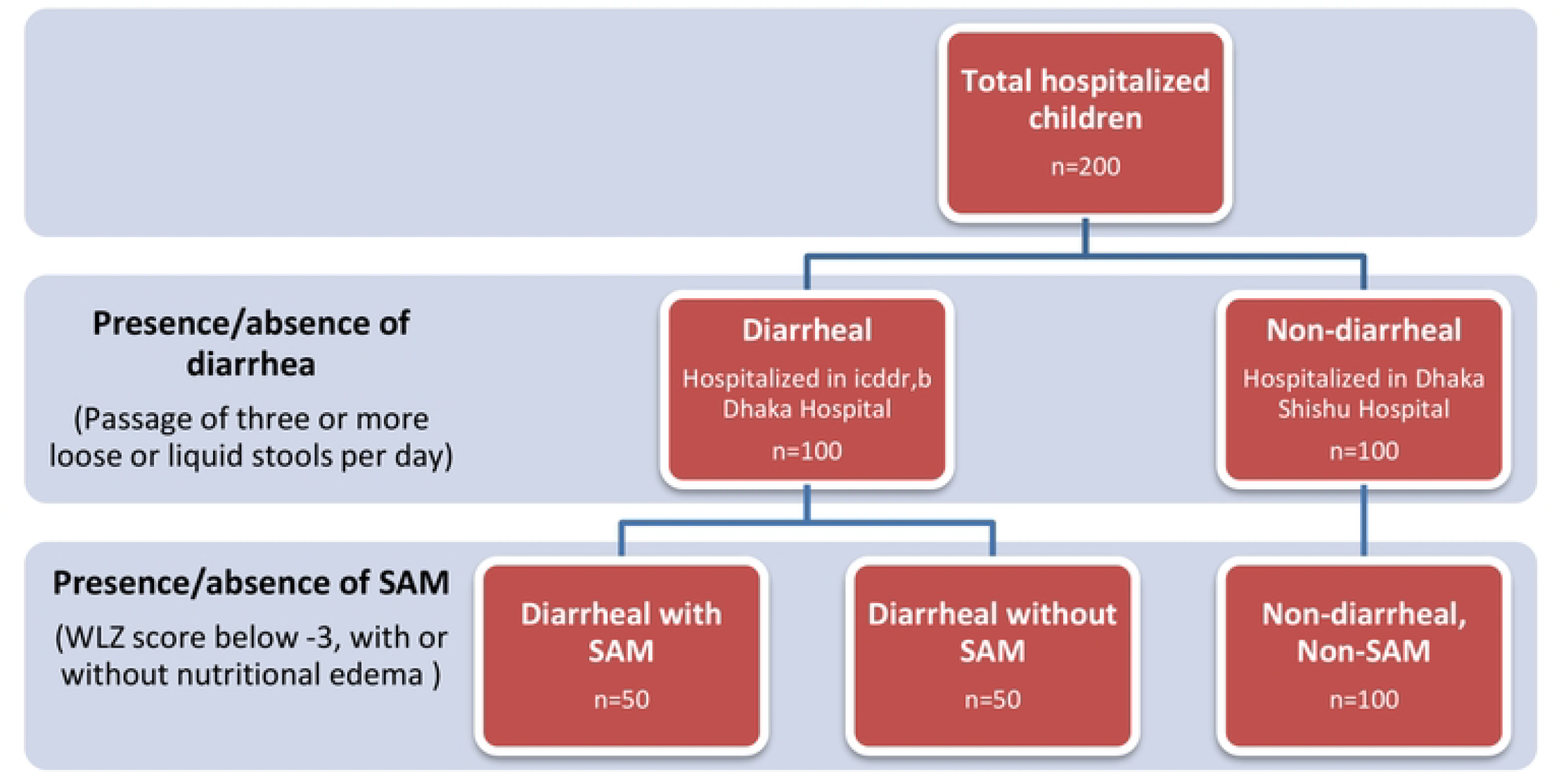
Categorization of study subjects

**Figure 3.**
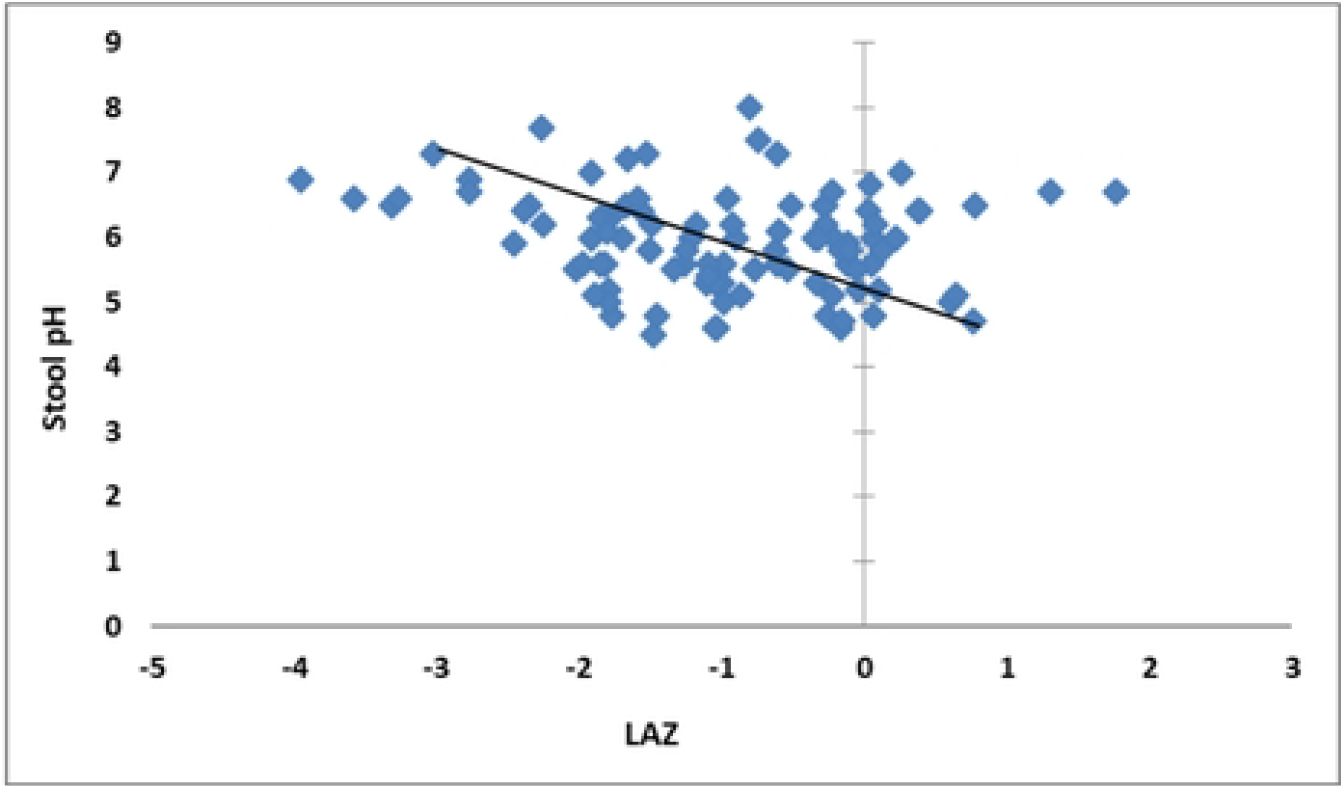
Scatter diagram showing negative correlation between fecal pH and LAZ scores in non-diarrheal children

Results of the bivariate linear regression with a significance level at or below 0.05 were included in the multivariate regression model. A statistically significant inverse association was observed between fecal pH with LAZ score of the non-diarrheal children (Table 2). A single unit increase in fecal pH was associated with 0.39 unit decrease in the LAZ score of the child. The other factors that showed a significant association with LAZ after the multivariate regression in non-diarrheal children were age, birth weight and mother’s height.

**Table 2.** Association of LAZ scores with fecal pH in non-diarrheal children. **Non-diarrheal cohort**, n=100 Outcome variable: LAZ

| Variables | Unadjusted |  |  | Adjusted |  |  |
| --- | --- | --- | --- | --- | --- | --- |
|  | B* | SE | P-value* | B* | SE | P-value* |
| Age | -0.002 | 0.001 | **0.006** | -0.002 | 0.001 | **0.041** |
| Sex | 0.331 | 0.232 | 0.157 |  |  |  |
| Birth weight | 0.612 | 0.228 | **0.009** | 0.616 | 0.202 | **0.003** |
| Mother's height | 0.076 | 0.021 | **0.001** | 0.077 | 0.021 | **0.000** |
| Mother's weight | 0.003 | 0.010 | 0.798 |  |  |  |
| Duration of illness | -0.001 | 0.011 | 0.919 |  |  |  |
| Duration of hospital stay | -0.99 | 0.050 | 0.052 |  |  |  |
| Antibiotics received | -1.643 | 1.066 | 0.126 |  |  |  |
| Stool pH | -0.300 | 0.139 | **0.034** | -0.389 | .142 | **0.008** |
| Mode of delivery | -0.148 | 0.232 | 0.525 |  |  |  |
| Duration of EBF | -0.001 | 0.002 | 0.558 |  |  |  |
| Currently breast fed | 0.155 | 0.268 | 0.565 |  |  |  |
| MDD* | -0.283 | 0.216 | 0.194 |  |  |  |
| WAMI* | 1.184 | 0.717 | 0.102 |  |  |  |

\*B=Regression co-efficient; p-value = Significance level; MDD=Minimum dietary diversity; WAMI= incorporation of several aspects of socio-economic status: access to improved Water and sanitation, the selected approach to measuring household wealth (Assets), Maternal education, and Income

Results of the bivariate linear regression in case of the diarrheal children revealed no association between fecal pH with LAZ score of the children (Table 3). However, age, mothers’s height and weight, antibiotic intake and duration of hospital stay were found to have significant relation in the bivariate analysis. The factors that showed a significant association with LAZ after the multivariate regression in diarrheal children were age, mother’s height, duration of hospital stay and receiving of antibiotics.

**Table 3.** Association of LAZ scores with fecal pH in diarrheal children. **Diarrheal cohort**, n=100 Outcome variable: LAZ

| Variables | Unadjusted |  |  | Adjusted |  |  |
| --- | --- | --- | --- | --- | --- | --- |
|  | B | SE | P-value | B | SE | P-value |
| Age | -0.004 | 0.001 | **0.000** | -0.003 | 0.001 | **0.002** |
| Sex | 0.468 | 0.265 | 0.081 |  |  |  |
| Mother's height | 0.064 | 0.023 | **0.006** | 0.058 | 0.021 | **0.008** |
| Mother's weight | 0.029 | 0.012 | **0.020** | 0.000 | 0.011 | 0.989 |
| Duration of illness | -0.015 | 0.033 | 0.652 |  |  |  |
| Duration of hospital stay | -0.235 | 0.058 | **0.000** | -0.209 | 0.054 | **0.000** |
| Antibiotics received | -0.957 | 0.276 | **0.001** | -0.700 | 0.246 | **0.005** |
| Stool pH | -0.020 | 0.131 | 0.880 |  |  |  |
| Mode of delivery | -0.245 | 0.252 | 0.335 |  |  |  |
| Duration of EBF | 0.001 | 0.002 | 0.992 |  |  |  |
| Currently breast fed | -0.140 | 0.296 | 0.637 |  |  |  |
| MDD | -0.074 | 0.422 | 0.862 |  |  |  |
| WAMI | 0.568 | 0.733 | 0.440 |  |  |  |

## Discussion

Our study showed that, elevated fecal pH in non-diarrheal children has a significant association with stunted growth. This finding is consistent with the findings of a previous study conducted among stunted and at risk of stunting children in the Mirpur slum community (15). Previous studies have already established an association of fecal pH with severe acute malnutrition (SAM) (30). Now, a relationship with chronic malnutrition is also observed through these studies.

Over the past century, there has been a tremendous advancement in medical science. This includes discovery of life-saving antibiotics, surgical procedures like the caesarian section and development of human milk replacers. All of these, however, had an impact on the gut microbiota. For instance, the practice of replacing human milk with infant formula is now widely observed. Unfortunately, many of such formulas lack the bacterial selectivity of human milk (6). Similarly, normal vaginal delivery is now broadly replaced by caesarean section delivery. Again, caesarian section restricts the natural transfer of microbiota from mother to the offspring (32). And there is also this overuse of antibiotics which is negatively affecting the gut microbial acquisition by the infant (33). Interestingly, there is reportedly an increasing trend of fecal pH that is also observed during this century. Studies demonstrated an increase of fecal pH in infants ranging from 5.0 to 6.5 between the period of 1926 to 2017 (6). This increase corresponds with a concurrent decrease of certain important gut microflora. *Bifidobacteria* and butyrate producing commensal *Clostridia* are such two very important microbiota that are present in the infant gut. Studies show, these microbiota contribute to formation of acidic feces, and an increased fecal pH denotes to decrease of these microflora in the intestine (6, 24, 34). A study conducted in Guatemala shows that both *Bifidobacteria* and *Clostridia* affects childhood stunting in a biologically plausible manner (35).

Gut microbiota has multiple protective functions on the gut, critical to optimal nutrient absorption (11, 36–38). However, there is limited information on gut microbiota of children in their first two years of life, a period when the maturation of microbiota is most essential (14). There is evidence that children in the community in discussion are breastfed along with complimentary feeding even beyond the first 2 years of age, creating a favorable environment in the gut for survival of *Bifidobacterium* up to this age (39). As reported by Bangladesh Demographic and Health Survey (BDHS) 2004, average duration of breastfeeding in Bangladeshi children was 31.9 months according to the findings of a study (39). *Bifidobacterium*, especially *Bifidobacterium longum* subsp. *Infantis* (*B. infantis*) has the unique ability to consume the full range of human milk oligosachharides (HMOs) (40). HMOs are beneficial in two ways - the first is that HMOs themselves act as nutrients for colon bacteria, and the second is that HMOs promote growth of nonpathogenic microflora preventing pathogenic microorganisms from adhesion to the host’s epithelial surface (16). The end product of the process are acidic in nature resulting in a meaningful effect on fecal pH (6).

*Clostridia* clusters XIVa and IV constitutes up to 10%-40% of the total bacterial population of the gut microbial community (24) and induces colonic T regulatory cell (Treg) accumulation (41), which strengthens pediatric gut immunity (22). As already discussed, butyrate released by these commensals through their metabolic functions lower the colonic pH and contributes to stool acidification (24). Abundance of SCFAs in the gut, particularly butyrate, is considered protective as they are calorie-rich nutrients which act in multiple ways to prevent undernutrition and stunting (22). SCFAs act as energy source for colonocytes, host metabolism regulators, and prevents the colonization of both aerobic and facultative anaerobic enteropathogens reducing the frequency and severity of GI diseases (22, 42).

However, it was also observed that such association does not apply for diarrheal stool samples. This is perhaps due to the pre-existing knowledge on reduced pH in diarrheal samples, which might have interfered. Studies show, *E*.*coli* and rotaviral diarrhea, amoebic dysentery, and severe acute malnutrition (SAM) often accompanied by carbohydrate (CHO) intolerance is significantly associated with decreased fecal pH (2, 30). Keeping this in mind, assessment of fecal pH in diarrheal and non-diarrheal fecal samples, keeping both in the same balance, was not deemed justified at the first place and was not attempted as well. However, a trend associating fecal pH and linear growth was expected, which was, however, not observed.

As per the discussion above, there is sufficient evidence on the positive roles of certain gut microflora, specifically *Bifidobacteria* and commensal *Clostridia* on the growth of children (13, 14). Characterization of fecal microbiome is an expensive process. As per the findings of this study, fecal pH is an indirect indicator of the abundance of these protective species in the gut of non-diarrheal children. As the method is very useful, non-invasive, inexpensive and above all can be conducted bedside with zero chance of any adverse events, it might be considered as a novel method of determination of stunted growth in early childhood.

## Limitations

Characterization of fecal microbiome of the children was not possible to be explored due to budgetary constraints, which would have generated important findings. Validation of this method with actual characterization of fecal microbiome is warranted, which remained a major limitation of this study.

## Conclusion

Elevated fecal pH in non-diarrheal children had significant association with childhood stunting, which was however not the case in diarrheal children. Fecal pH might be a possible indirect determinant of stunting in non-diarrheal children. Further studies exploring characterization of the fecal microbiome are warranted to validate this finding.

## Data Availability

All relevant data are within the manuscript and its Supporting Information files.

## 3 Author Contributions

TA originated the idea for the study. TA and MSH participated in the design of the study. MSH, MMH, and TA conducted the study. MSH supervised the sample and data collection and sample analysis. MSH, MM and TA were involved in data analysis. MSH and TA interpreted the results. MSH, MMH, MM and TA were involved in manuscript writing. All authors read and approved the final manuscript.

## 5 Acknowledgments

This work was supported, in whole, by the Gates Foundation [OPP1136751]. The conclusions and opinions expressed in this work are those of the author(s) alone and shall not be attributed to the Foundation. Under the grant conditions of the Foundation, a Creative Commons Attribution 4.0 License has already been assigned to the Author Accepted Manuscript version that might arise from this submission. Please note works submitted as a preprint have not undergone a peer review process. icddr,b acknowledges with gratitude the commitment of the Gates Foundation to its research efforts. icddr,b also gratefully acknowledges the following donors who provide unrestricted support: Government of the People’s Republic of Bangladesh and Global Affairs Canada (GAC).

## 6 Data Availability Statement

The raw data supporting the conclusions of this article will be made available by the authors on request, without undue reservation.

